# Genetic and Lifestyle Risks for Coronary Artery Disease and Long-Term Risk of Incident Dementia Subtypes

**DOI:** 10.1101/2024.07.17.24310606

**Authors:** Arisa Sittichokkananon, Victoria Garfield, Scott T. Chiesa

**Affiliations:** Princess Srisavangavadhana College of Medicine; Chulabhorn Royal Academy; Bangkok; Thailand; Department of Pharmacology & Therapeutics; Institute of Systems, Molecular, & Integrative Biology; University of Liverpool, Liverpool, UK; Medical Research Council Unit for Lifelong Health and Ageing at UCL; Institute of Cardiovascular Science; UCL; London; UK

## Abstract

**Background:** Shared genetic and lifestyle risk factors may underlie the development of both coronary artery disease (CAD) and dementia. This study aimed to examine if an increased genetic risk for CAD is associated with long-term risk of developing all-cause, Alzheimer’s, or vascular dementia, and investigate whether the presence of healthy lifestyle behaviours in the mid-to-late life period may attenuate this risk.

**Methods:** A prospective cohort study of 365,782 participants free from dementia for at least 5 years post-baseline assessment was conducted within the UK Biobank study. Genetic risk was assessed using a genome-wide polygenic risk score (PRS) for CAD, and lifestyle risk using a modified version of the American Heart Association’s Life’s Essential 8 Lifestyle Risk Score (LRS). Primary outcomes were incident all-cause, Alzheimer’s, and vascular dementia diagnoses obtained from linked electronic health records. Secondary outcomes were neuroimaging phenotypes with well-established links to future dementia risk measured in 32,592 participants recalled for MRI imaging.

**Results:** 8,870 cases of all-cause dementia were observed over a median 13.9-year follow-up. Higher genetic risk for CAD was associated with an elevated risk of all dementia subtypes (HRs = 1.08-1.16; p<0.001 for all). A higher LRS was associated with a modestly increased risk of all-cause dementia (HR = 1.06 [1.04-1.08]; p < 0.001), with this risk likely arising through increased rates of vascular dementia (HR = 1.22 [1.17-1.28]) as no evidence was found for any associations with Alzheimer’s disease (HR = 0.99 [0.95-1.02]; p = 0.535). Individuals with a combination of high genetic and high lifestyle risk scores for CAD were more than twice as likely to develop vascular dementia during long-term follow-up compared to those with low levels of both. This risk was substantially attenuated in those following healthy lifestyle behaviours at baseline, however, regardless of underlying genetic risk (e.g. HR for low vs high lifestyle risk scores = 1.43 [1.12-1.81] vs. 2.16 [1.73-2.69] respectively in individuals with high genetic risk). In a subset of individuals recalled for neuroimaging assessments, those with high genetic and lifestyle risk for CAD demonstrated a 30% greater volume of white matter hyperintensities than those with low risk, while showing little difference in grey matter or hippocampal volumes.

**Conclusions:** Individuals who are genetically predisposed to developing CAD also face an increased risk of developing dementia in old age. This risk is reduced in those adopting healthy lifestyle behaviours earlier in the lifespan, however, particularly in those at risk from dementia caused by underlying vascular pathology.

## INTRODUCTION

Coronary artery disease (CAD) and dementia are two of the leading causes of death and disability worldwide, accounting for an estimated annual burden of 9 million and 2 million deaths, respectively (1,2). While both conditions occur as the result of separate and complex disease processes arising from diverse aetiologies, accumulating evidence suggests that each may share common – and at times potentially modifiable – underlying risk factors that act to simultaneously increase the risk of both diseases.

The most common cause of dementia is Alzheimer’s Disease (AD), responsible for 60-70% of dementia diagnoses (3). AD is characterized by a progressive decline in memory and thinking skills that are believed to arise from an accumulation of amyloid plaques and neurofibrillary tau tangles in the brain that are the hallmark of a diagnosis. Recent years have seen a wealth of research investigating the role that genetic variants play in the development of AD (4–6), and how this risk may be attenuated by the adoption of healthy lifestyle behaviours at a younger age (7–10).

Cerebrovascular disease also contributes to a significant proportion of dementia diagnoses, however, both in the form of overt vascular dementia and as a co-pathology present in an estimated 50-80% of Alzheimer’s diagnoses (11–13) As atherosclerosis-related hypoperfusion plays a major role in both CAD and cerebrovascular disease (14,15), an underlying genetic predisposition to atherosclerosis may therefore represent an important link in the relationship between heart and brain health. In support of this, polygenic risk scores (PRS) for atherosclerotic CAD have previously been linked to both reduced cognitive function (16) and brain atrophy (17) in later life, implying that shared genetic pathways underlying CAD may also increase risk of future dementia. No study to-date has directly tested this relationship over long-term follow-up or investigated the impact that healthy lifestyle behaviours earlier in the lifespan may have on different dementia outcomes.

Utilising data from a large-scale cohort study of over 360,000 participants prospectively followed for 14 years, our primary aims were to 1) assess whether a PRS for CAD was associated with an increased risk of developing three common dementia subtypes (all-cause, Alzheimer’s Disease, and vascular) during the transition through mid-to-late life, and 2) assess whether this risk was attenuated in those following healthy lifestyle behaviours at their baseline assessment. A secondary aim was to assess underlying changes in brain structure that may explain these differences in risk in a subset of over 35,000 individuals recalled for MRI neuroimaging.

## METHODS

### Data Availability

All data used in this publication are open access and available to *bona fide* researchers through well-documented processes detailed at https://www.ukbiobank.ac.uk/. The statistical code for all analyses in this paper can be found in an open-access GitHub repository located at https://github.com/scottchiesa/UKB_PRS_LRS_Dementia.

### Study Population

This study used data from the UK Biobank, a large population-based cohort of over 500,000 individuals recruited in the United Kingdom during 2006-2010. The UK Biobank study received ethical approval from the National Health Service North-West Multicentre Research Ethics Committee. All participants gave informed consent during the baseline assessment. Any participants withdrawing consent prior to commencement of this study were excluded before analysis. Participants were also excluded if they had prevalent dementia (a diagnosis of dementia prior to baseline), were diagnosed with dementia within 5 years of baseline (to minimise risk of undiagnosed dementia cases), or were younger than 50 years at baseline (as the primary outcome of interest in this study was late-onset dementia, which occurs after age 65). Participants with non-White ancestry were also excluded to minimise residual confounding through underlying ancestral differences and possible population stratification (18).

### Exposures

#### Polygenic Risk Score (PRS)

This study used the standard UK Biobank PRS for CAD derived by Thompson et al, full details of which are described elsewhere (19). In brief, a Bayesian approach was used to generate the CAD PRS from a meta-analysis of GWAS summary statistics using multiple external studies. Per-individual PRS values were calculated as the genome-wide sum of the per-variant posterior effect size multiplied by allele dosage. To avoid the risk of overfitting, the PRS was developed based only on non-UK-Biobank populations before being calculated for all individuals in UK Biobank.

#### Lifestyle Risk Score (LRS)

A lifestyle risk score (LRS) was next created based on an adapted form of the American Heart Association’s “Life’s Essential 8 (LE8)” concept, created to measure and monitor cardiovascular health. LE8 consists of 8 components: diet, physical activity, smoking status, sleep duration, BMI, blood lipids, blood glucose, and blood pressure (20). Physical activity, smoking status, sleep quality, and diet were assessed at the initial assessment centre visit using a touchscreen questionnaire. Participants were asked whether they currently smoke or if they have ever smoked in the past. They were also asked about the frequency, duration, and intensity of their physical activities. Sleep quality was assessed by self-reporting how many hours of sleep they got in every 24 hours. For diet, participants answered questions about how many servings of fruit, vegetables, whole grains, and refined grains they had each day, and how many servings of processed meats, unprocessed red meats, and fish they had each week. BMI values were calculated as weight(kg) / height(m)^2^ from height and weight measurements taken at the baseline visit. Blood pressure was reported as the average of two automated measures taken a few moments apart using an Omron device at baseline visit. For biomarkers, blood samples were collected from participants at each assessment centre, refrigerated, and transported to a central laboratory for automated processing. LDL-C was measured by enzymatic protective selection analysis on a Beckman Coulter AU580. HbA1c was used in place of fasting glucose to provide a better long-term marker of glycaemic status and was measured using high-performance liquid chromatography analysis on a Bio-Rad VARIANT II Turbo. Extreme values +/- 4 standard deviations (SD) from the mean were excluded from the LRS calculation. Participants were then scored 0-2 points for each lifestyle component, with higher scores representing greater cardiovascular risk (Table 1). The points from each lifestyle category were then summed to give an overall risk score of 0-16, with higher numbers indicating greater risk.

**Table 1:**
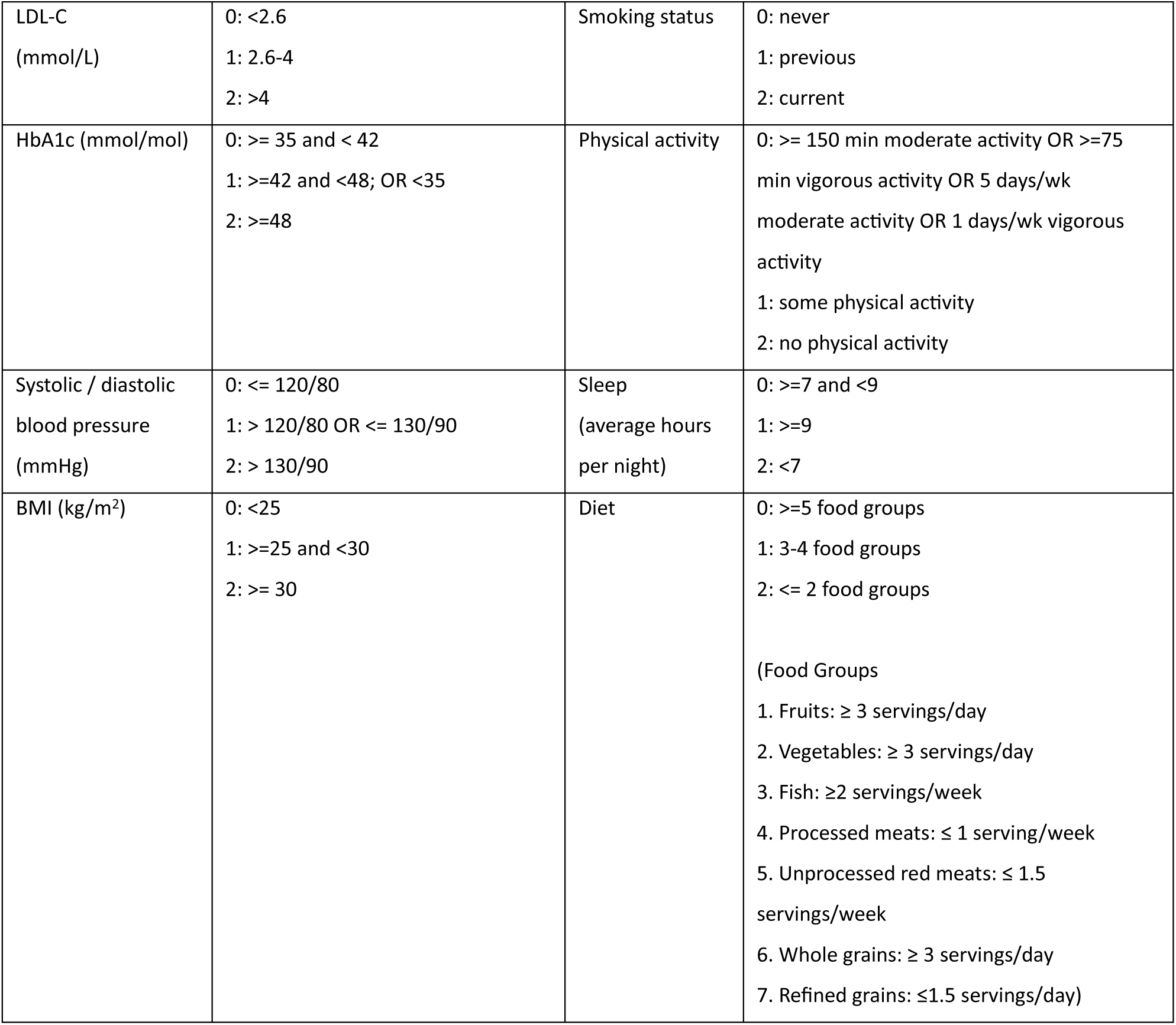
Definition of Cardiovascular Health Metrics. BMI, body mass index; BP, blood pressure; HbA1, glycated haemoglobin; LDL-C, low-density lipoprotein cholesterol.

|  |  |  |  |
| --- | --- | --- | --- |
| LDL-C<br>(mmol/L) | 0: <2.6<br>1: 2.6-4<br>2: >4 | Smoking status | 0: never<br>1: previous<br>2: current |
| HbA1c (mmol/mol) | 0: $\geq 35$ and < 42<br>1: $\geq 42$ and <48; OR <35<br>2: $\geq 48$ | Physical activity | 0: $\geq 150$ min moderate activity OR $\geq 75$ min vigorous activity OR 5 days/wk moderate activity OR 1 days/wk vigorous activity<br>1: some physical activity<br>2: no physical activity |
| Systolic / diastolic blood pressure<br>(mmHg) | 0: $\leq 120/80$<br>1: > 120/80 OR $\leq 130/90$<br>2: > 130/90 | Sleep<br>(average hours per night) | 0: $\geq 7$ and <9<br>1: $\geq 9$<br>2: <7 |
| BMI (kg/m<sup>2</sup>) | 0: <25<br>1: $\geq 25$ and <30<br>2: $\geq 30$ | Diet | 0: $\geq 5$ food groups<br>1: 3-4 food groups<br>2: $\leq 2$ food groups<br>(Food Groups<br>1. Fruits: $\geq 3$ servings/day<br>2. Vegetables: $\geq 3$ servings/day<br>3. Fish: $\geq 2$ servings/week<br>4. Processed meats: $\leq 1$ serving/week<br>5. Unprocessed red meats: $\leq 1.5$ servings/week<br>6. Whole grains: $\geq 3$ servings/day<br>7. Refined grains: $\leq 1.5$ servings/day) |

### Outcomes

#### Dementia Diagnoses

International Classification of Diseases (ICD)-10 codes were used to identify dementia cases in linked hospital episode statistics data. Incident dementia was captured using the algorithmic method detailed in Wilkinson et al (21), which used data from linked UK hospital admission, primary care, and mortality records. Three subtypes of dementia were included in this analysis: all-cause dementia, Alzheimer’s disease, and vascular dementia.

#### Neuroimaging Phenotypes

Three common neuroimaging phenotypes with well-established links to future risk of dementia were chosen as imaging outcomes in secondary analyses: cortical grey matter volume (22), hippocampal volume (23), and white matter hyperintensity volume (24). Full details on the imaging-derived phenotypes available in UK Biobank can be found at (25). Briefly, however, grey matter volume was extracted from T1-weighted images using the FMRIB Automated Segmentation Tool (FAST; version 4.1) and hippocampal volume using the FMRIB Integrated Registration and Segmentation Tool (FIRST; version 5.0). White matter hyperintensity volumes were calculated based on T1 and T2 FLAIR, and derived using the Brain Intensity Abnormality Classification Algorithm (BIANCA) (26).

### Covariates

Covariates included age, sex, level of education completed (below secondary / secondary / or higher), and socioeconomic status (Townsend deprivation index). Age was included as a covariate as dementia prevalence substantially increases with age (27). Sex was considered as a covariate as the ratio of male to female prevalence is different in each dementia subtype, with Alzheimer’s disease being more common in women and vascular dementia being more prevalent in men (28). Increased educational attainment is associated with reduced risk of both CAD risk factors and dementia (29), while lower socioeconomic status is associated with a greater risk (30). For neuroimaging analyses, grey matter and hippocampal volumes were additionally adjusted for total brain volume and childhood body size due to the known risk of confounding from these factors (31).

### Statistical Analyses

Descriptive characteristics of the cohort were summarized by dementia status using percentages for categorical variables, means and SDs for normally distributed continuous variables, and median and IQR for non-normally distributed variables. Normality was assessed via visual inspection of histograms. LRS was found to be normally distributed and was therefore z-scored prior to inclusion in models to enable comparability to PRS models.

#### Dementia Diagnoses

Cox proportional hazards models were used to examine associations between PRS and LRS as exposures and all-cause dementia, Alzheimer’s disease, and vascular dementia as outcomes. Interactions between PRS and LRS were also tested, as were interactions between both exposures and sex. As the latter showed some evidence for a modifying effect of sex on multiple outcomes, all Cox models were run with sex included as a stratified variable, providing equal coefficients across strata in the pooled data while allowing a distinct baseline hazard for each sex. Time scale of follow-up was time since baseline assessment (2006-2010) until 13^th^ December 2022. The proportionality of hazards assumption for each model was assessed using the Schoenfeld residuals technique (32). For PRS exposures, statistical models were adjusted for age and sex alone. For LRS exposures, models were additionally adjusted for highest education level and socioeconomic status due to their known associations with lifestyle health factors. The decision not to adjust PRS models for education and socioeconomic status was taken to avoid overadjustment bias due to the possibility that these factors may lie on the causal pathway linking genetic risk for CAD to dementia development, and thus may represent effect mediators rather than confounders. To examine incident dementia risk according to combined genetic and lifestyle risk, PRS and LRS were each stratified into categories before being combined for analyses. PRS was split into 3 equal tertiles, where 33.3% of participants had low, intermediate, and high genetic risk, respectively. LRS was also split into 3 approximately equal groups, with 34.4% of participants categorized as having low lifestyle risk (i.e., scoring <6 points), 35.4% having intermediate lifestyle risk (i.e., scoring 6-7 points), and 30.2% having high lifestyle risk (i.e., scoring 8-16 points). Cox proportional hazards models were then performed on the 9 combined genetic and lifestyle risk categories, with combined low genetic and lifestyle risk as the reference group.

#### Neuroimaging Phenotypes

For neuroimaging measures, multivariable linear regression models were used to test associations between all of the exposures listed above and three structural neuroimaging outcomes; namely cortical grey matter volume, hippocampal volume, and white matter hyperintensity (WMH) volume. For ease of comparison, all outcomes were log-transformed prior to inclusion in models before being back-transformed for reporting, and are therefore expressed as a percentage change in geometric mean per unit change in exposure. PRS analyses were adjusted for age, sex, total brain volume (latter not for WMH) and childhood body size, while LRS analyses were adjusted for age, sex, highest education, current socioeconomic status, total brain volume (again not for WMH) and childhood body size.

#### Statistical Interpretation and Sensitivity Analyses

All analyses were performed in Stata Version 17 (College Station, TX: StataCorp LLC). Multiple imputation using chained equations (10 imputations) was used to account for missing data and two sensitivity analyses were additionally run for comparison with main results: 1) PRS and LRS models re-run with unimputed data, 2) PRS and LRS models re-run after exclusion of participants with evidence of an opposing subtype of dementia (e.g. removing people with a known vascular dementia diagnosis in analyses comparing people with and without an Alzheimer’s diagnosis). An *a-priori* decision was made to interpret findings mainly on the basis of model estimates and their 95% CIs rather than assign significance using an arbitrary p-value cut-off of 0.05 (33). These p-values are still highlighted throughout, however, for reference.

## RESULTS

### Sample Characteristics

The final study sample consisted of 365,782 individuals, 8,870 of whom had all-cause dementia during a median follow-up of 13.9 years (Supplemental Figure 1). Both PRS and LRS were normally distributed (Figure 1). Participants with dementia were on average older than participants without dementia. A greater proportion of participants with Alzheimer’s disease were female, while participants with vascular dementia were more likely to be male. Full baseline characteristics are presented in Table 2 and details on data missingness can be found in Supplemental Table 1.

**Figure 1:**
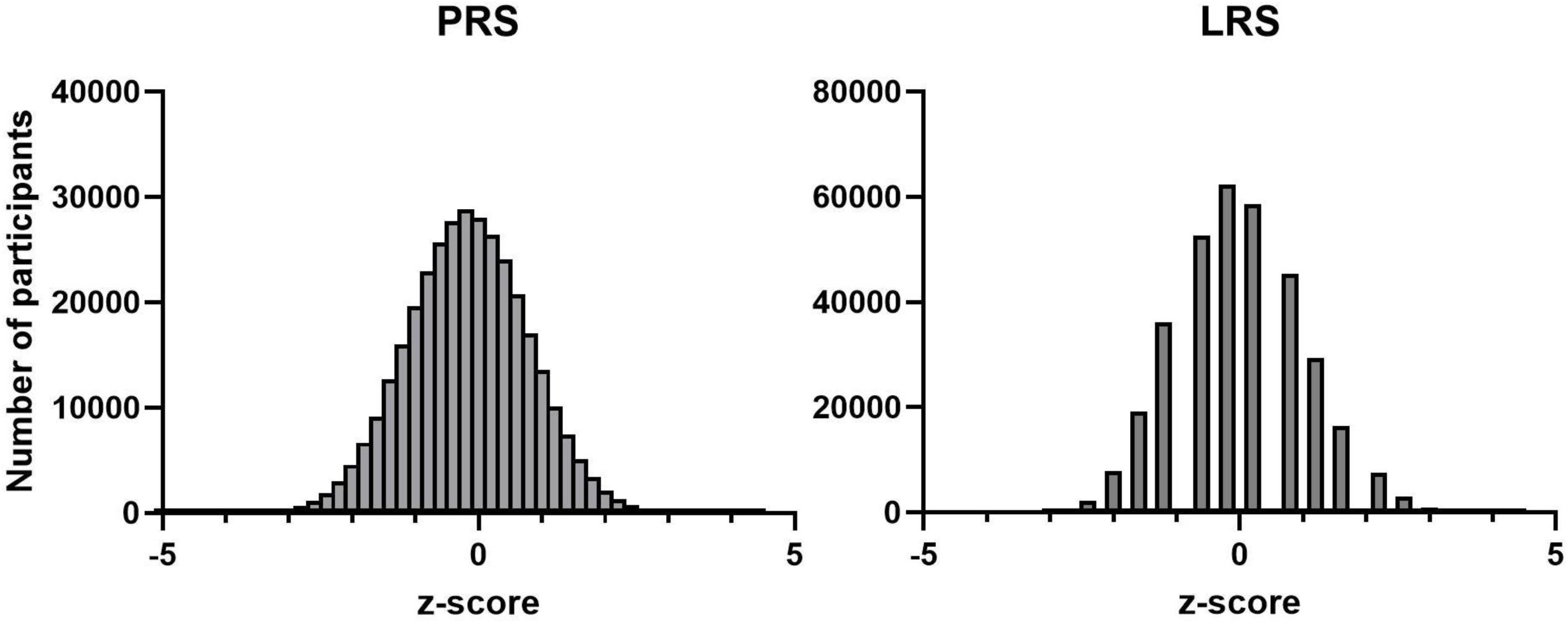
Distribution of Polygenic and Lifestyle Risk Scores for Coronary Artery Disease. PRS, polygenic risk score; LRS, lifestyle risk score.

**Figure 2:**
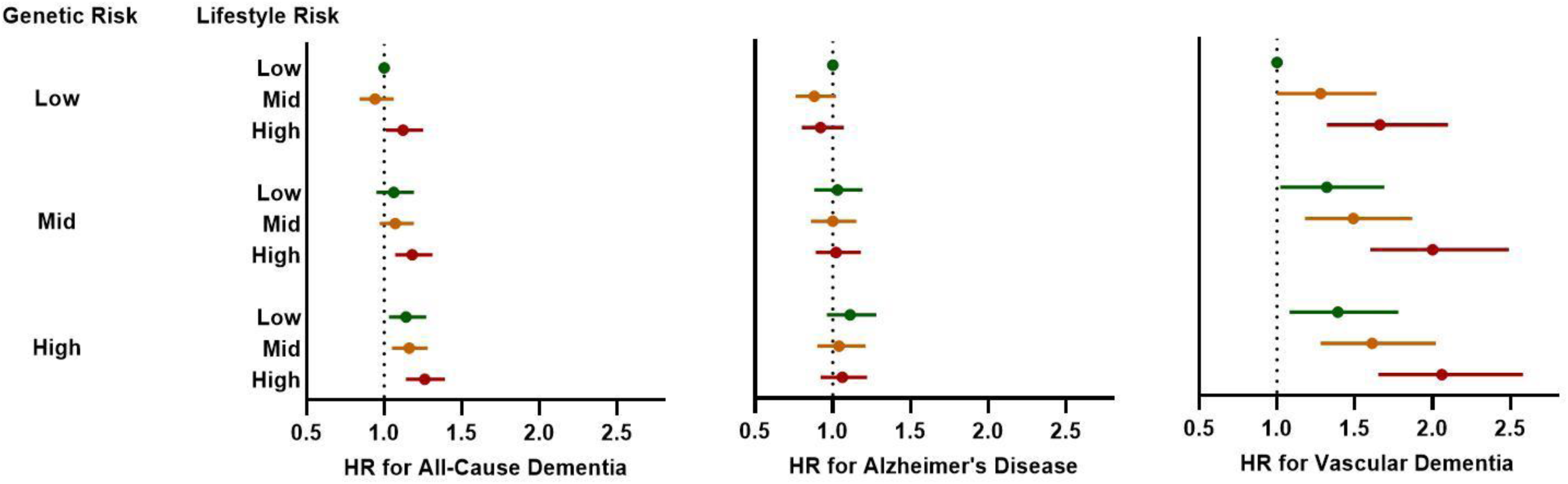
Forest Plots Showing Hazard Ratios for Combined Genetic and Lifestyle Risk for Coronary Artery Disease and Incident Dementia Subtypes. Cox regression models adjusting for age, sex, highest education level, and socioeconomic status. Data expressed as hazard ratios and 95% confidence intervals compared to reference category of combined low genetic and lifestyle risk. Low, mid, and high categories of genetic and lifestyle risk refer to tertiles of PRS and LRS, respectively. HR, hazard ratio.

**Figure 3:**
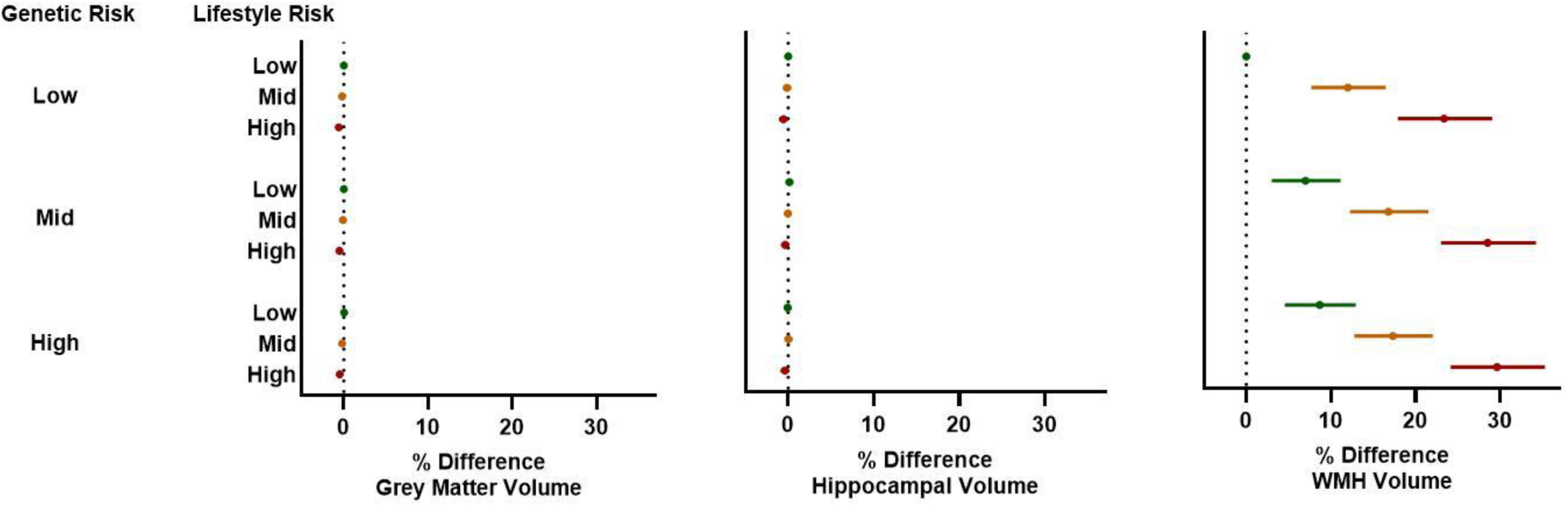
Forest Plots Showing Hazard Ratios for Combined Genetic and Lifestyle Risk for Coronary Artery Disease and Neuroimaging Phenotypes. Multivariable regression models adjusting for age, sex, highest education level, socioeconomic status, total brain volume (not WMH), and childhood body size. Data expressed as % difference in geometric mean and 95% confidence intervals compared to reference category of combined low genetic and lifestyle risk. Low, mid, and high categories of genetic and lifestyle risk refer to tertiles of PRS and LRS, respectively. HR, hazard ratio.

**Table 2:**
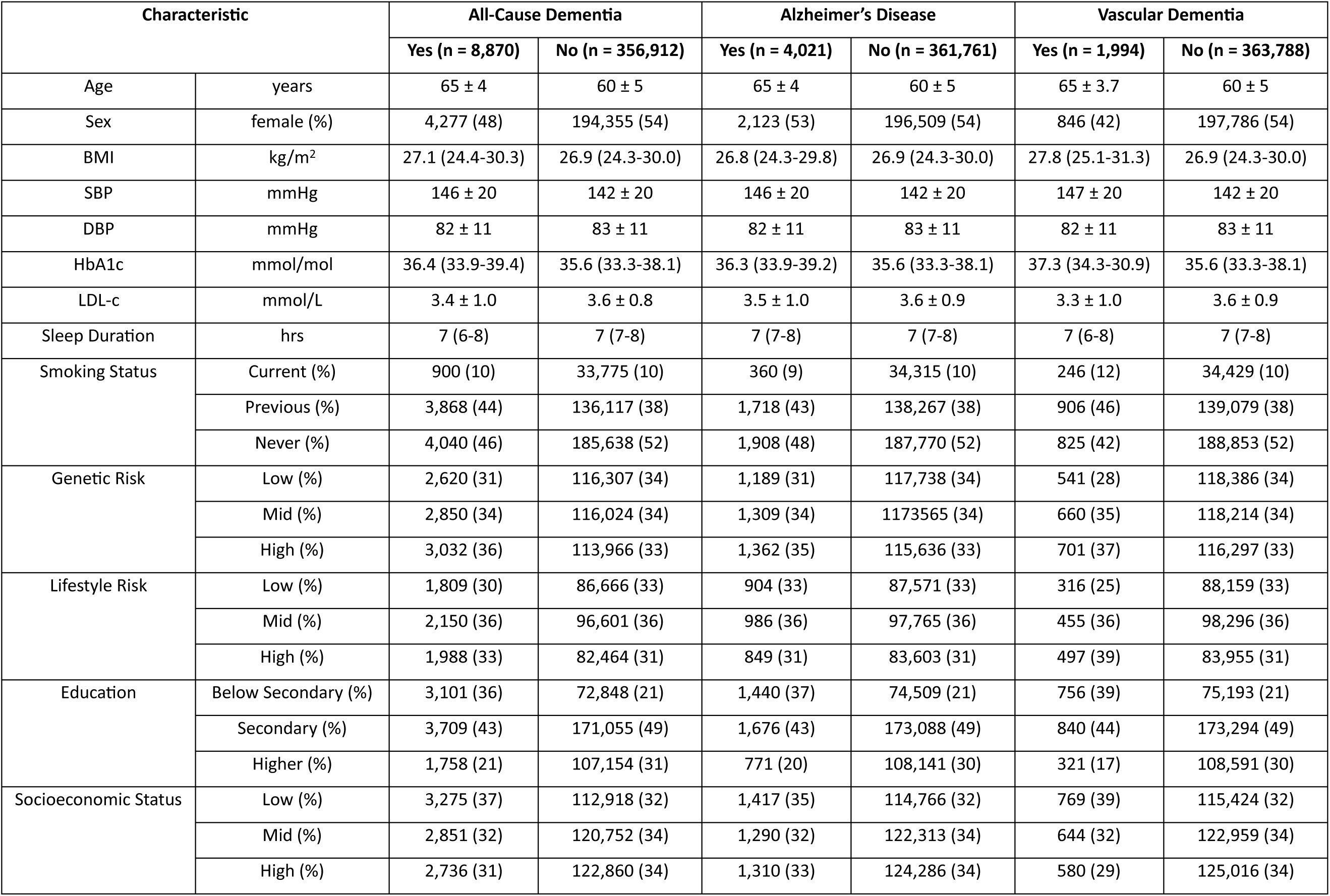
Participant Characteristics. Data represent mean ± SD, median (IQR), or n (%). BMI, body mass index; BP, blood pressure; HbA1, glycated haemoglobin; LDL-C, low-density lipoprotein cholesterol.

| Characteristic |  | All-Cause Dementia |  | Alzheimer's Disease |  | Vascular Dementia |  |
| --- | --- | --- | --- | --- | --- | --- | --- |
|  |  | Yes (n = 8,870) | No (n = 356,912) | Yes (n = 4,021) | No (n = 361,761) | Yes (n = 1,994) | No (n = 363,788) |
| Age | years | 65 ± 4 | 60 ± 5 | 65 ± 4 | 60 ± 5 | 65 ± 3.7 | 60 ± 5 |
| Sex | female (%) | 4,277 (48) | 194,355 (54) | 2,123 (53) | 196,509 (54) | 846 (42) | 197,786 (54) |
| BMI | kg/m<sup>2</sup> | 27.1 (24.4-30.3) | 26.9 (24.3-30.0) | 26.8 (24.3-29.8) | 26.9 (24.3-30.0) | 27.8 (25.1-31.3) | 26.9 (24.3-30.0) |
| SBP | mmHg | 146 ± 20 | 142 ± 20 | 146 ± 20 | 142 ± 20 | 147 ± 20 | 142 ± 20 |
| DBP | mmHg | 82 ± 11 | 83 ± 11 | 82 ± 11 | 83 ± 11 | 82 ± 11 | 83 ± 11 |
| HbA1c | mmol/mol | 36.4 (33.9-39.4) | 35.6 (33.3-38.1) | 36.3 (33.9-39.2) | 35.6 (33.3-38.1) | 37.3 (34.3-30.9) | 35.6 (33.3-38.1) |
| LDL-c | mmol/L | 3.4 ± 1.0 | 3.6 ± 0.8 | 3.5 ± 1.0 | 3.6 ± 0.9 | 3.3 ± 1.0 | 3.6 ± 0.9 |
| Sleep Duration | hrs | 7 (6-8) | 7 (7-8) | 7 (7-8) | 7 (7-8) | 7 (6-8) | 7 (7-8) |
| Smoking Status | Current (%) | 900 (10) | 33,775 (10) | 360 (9) | 34,315 (10) | 246 (12) | 34,429 (10) |
|  | Previous (%) | 3,868 (44) | 136,117 (38) | 1,718 (43) | 138,267 (38) | 906 (46) | 139,079 (38) |
|  | Never (%) | 4,040 (46) | 185,638 (52) | 1,908 (48) | 187,770 (52) | 825 (42) | 188,853 (52) |
| Genetic Risk | Low (%) | 2,620 (31) | 116,307 (34) | 1,189 (31) | 117,738 (34) | 541 (28) | 118,386 (34) |
|  | Mid (%) | 2,850 (34) | 116,024 (34) | 1,309 (34) | 117,356 (34) | 660 (35) | 118,214 (34) |
|  | High (%) | 3,032 (36) | 113,966 (33) | 1,362 (35) | 115,636 (33) | 701 (37) | 116,297 (33) |
| Lifestyle Risk | Low (%) | 1,809 (30) | 86,666 (33) | 904 (33) | 87,571 (33) | 316 (25) | 88,159 (33) |
|  | Mid (%) | 2,150 (36) | 96,601 (36) | 986 (36) | 97,765 (36) | 455 (36) | 98,296 (36) |
|  | High (%) | 1,988 (33) | 82,464 (31) | 849 (31) | 83,603 (31) | 497 (39) | 83,955 (31) |
| Education | Below Secondary (%) | 3,101 (36) | 72,848 (21) | 1,440 (37) | 74,509 (21) | 756 (39) | 75,193 (21) |
|  | Secondary (%) | 3,709 (43) | 171,055 (49) | 1,676 (43) | 173,088 (49) | 840 (44) | 173,294 (49) |
|  | Higher (%) | 1,758 (21) | 107,154 (31) | 771 (20) | 108,141 (30) | 321 (17) | 108,591 (30) |
| Socioeconomic Status | Low (%) | 3,275 (37) | 112,918 (32) | 1,417 (35) | 114,766 (32) | 769 (39) | 115,424 (32) |
|  | Mid (%) | 2,851 (32) | 120,752 (34) | 1,290 (32) | 122,313 (34) | 644 (32) | 122,959 (34) |
|  | High (%) | 2,736 (31) | 122,860 (34) | 1,310 (33) | 124,286 (34) | 580 (29) | 125,016 (34) |

### Genetic Risk for CAD and Future Dementia Risk

Higher genetic risk for CAD was associated with increased risk of developing all dementia subtypes, both when expressed as a continuous variable and when stratified by tertiles (Table 3). This risk appeared to be greatest for vascular dementia, with those in the top tertile for CAD PRS showing almost double the risk for a vascular diagnosis of dementia compared to Alzheimer’s disease (∼32% vs 17%, respectively; Table 3). Similar results were observed in sensitivity analyses (Supplemental Table 2).

**Table 3:**
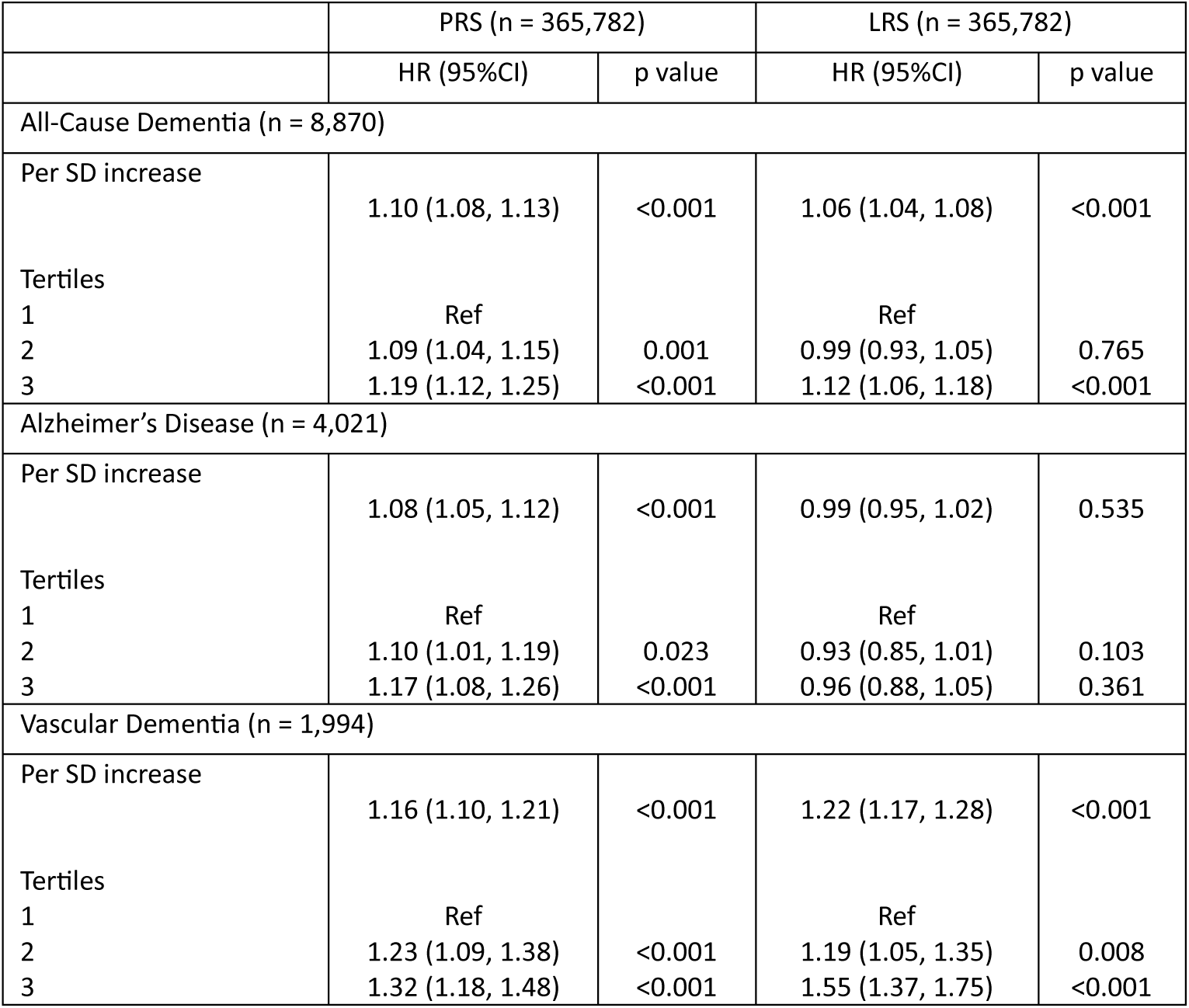
Associations Between Genetic and Lifestyle Risk for Coronary Artery Disease and Risk of Incident Dementia Subtypes. PRS, polygenic risk score; LRS, lifestyle risk score. PRS adjusted for age and sex. LRS adjusted for age, sex, highest education level, and current socioeconomic status.

### Lifestyle Risk for CAD and Future Dementia Risk

Higher lifestyle risk for CAD was associated with an increased risk of developing all-cause dementia (HR [95%CI] = 1.12 [1.06, 1.18]; p < 0.001; Table 3). This association appeared to be driven almost exclusively by vascular dementia, with those in the top tertile for CAD LRS having a > 50% increase in risk of vascular dementia in fully-adjusted models compared to those in the lowest tertile (HR [95%CI = 1.55 [1.37, 1.75]; p < 0.001; Table 3), whereas no association was seen for Alzheimer’s disease (HR [95%CI = 0.96 [0.88, 1.05]; p = 0.361; Table 3). Similar results were again observed in sensitivity analyses (Supplemental Table 2).

### Combined Genetic and Lifestyle Risk for CAD and Future Dementia Risk

There was little evidence for an interaction between PRS and LRS for either all-cause dementia (p=0.383), Alzheimer’s disease (p=0.917), or vascular dementia (p=0.793), suggesting that the association between lifestyle risk factors and future dementia risk did not vary depending on an individual’s underlying genetic risk. However, a combination of high genetic and lifestyle risk was found to have an additive effect on future dementia risk (Figure 5). This once again appeared predominantly attributable to associations between CAD risk factors and vascular dementia, with individuals in the highest tertile for both PRS and LRS found to have more than double the risk of developing vascular dementia during long-term follow-up than those in the lowest tertiles (HR = 2.16 [1.73-2.69]; p<0.001). In contrast, little evidence was found for any additive effect of genetic and lifestyle risks for CAD on Alzheimer’s disease (HR = 1.06 [0.93-1.22]; p=0.391).

### Genetic and Lifestyle Risk for CAD and Differences in Subclinical Neuroimaging Phenotypes

White matter hyperintensity volumes were associated with both genetic and lifestyle risks for CAD, with an increase in WMH volume of ∼3% and 9% observed for every SD increase in PRS and LRS, respectively (p<0.001 for both; Table 4). When combining genetic and lifestyle risks, individuals with risk scores in the top tertile for both PRS and LRS were found to have WMH volumes ∼30% higher than those with low levels of each (Figure 5). In contrast, no association was seen between the CAD PRS and either grey matter or hippocampal volumes (Table 4). While there was some evidence of an association between the LRS and these same outcomes, the absolute changes were clinically negligible (∼0.2% decrease in both volumes per SD increase in LRS; Table 4).

**Table 4:** Associations Between Genetic and Lifestyle Risks for Coronary Artery Disease and Subclinical Neuroimaging Phenotypes. PRS, polygenic risk score; LRS, lifestyle risk score; WMH, white matter hyperintensities. PRS adjusted for age, sex, and total brain volume (latter not for WMH). LRS adjusted for age, sex, total brain volume (not WMH), highest education, current socioeconomic status, and childhood body size.

|  |  | PRS |  | LRS |  |
| --- | --- | --- | --- | --- | --- |
|  | N | % Change in Outcome<br>per SD Increase in<br>Exposure (95% CI) | p<br>value | % Change in Outcome<br>per SD Increase in<br>Exposure (95% CI) | p<br>value |
| Grey Matter<br>Volume | 32,592 | 0.0 (0.0, 0.0) | 0.852 | -0.2 (-0.3, -0.2) | <0.001 |
| Hippocampal<br>Volume | 32,578 | 0.0 (-0.1, 0.1) | 0.840 | -0.2 (-0.3, -0.1) | 0.002 |
| WMH<br>Volume | 31,623 | 3.4 (2.3, 4.5) | <0.001 | 9.4 (8.2, 10.5) | <0.001 |

## DISCUSSION

In this large UK-based longitudinal population study followed for up to 14 years, we provide four novel and important findings linking heart and brain health. First, we show that individuals who are genetically predisposed to developing coronary artery disease also face an increased risk of developing dementia in later-life. Second, we demonstrate that this risk is reduced in those maintaining good cardiovascular health in the years preceding diagnosis, regardless of underlying genetic predisposition. Third, we show that this lifestyle-related risk reduction appears to be driven almost solely by a reduced incidence of vascular dementia cases, rather than through any association with Alzheimer’s disease. Finally, in a sub-set of individuals free from dementia at time of assessment, we provide evidence for early signs of vascular damage (i.e. white matter hyperintensity burden) that broadly mirror patterns observed for progression to vascular dementia.

While associations between genetic variation and dementia risk have been extensively studied in recent years, these have almost exclusively been for Alzheimer’s disease – the most common underlying cause of dementia (7,8,34). To the best of our knowledge, no study to-date has addressed the impact that a genetic predisposition to atherosclerotic vascular disease may also have on future dementia risk. Given well-established links in the literature between heart and brain health (35), alongside prior evidence for reductions in cognitive (16) and brain reserve (17) in individuals at high genetic risk for coronary heart disease, we hypothesized that genetic variants associated with an increased risk for CAD may also increase the risk of dementia in those surviving to old age. Our findings support this hypothesis, demonstrating a ∼20% increased risk of all-cause dementia when comparing individuals in the top vs bottom tertile for a genome-wide CAD PRS. By utilizing the statistical power afforded by the large number of incident dementia cases available in UK Biobank to separate these diagnoses into Alzheimer’s and vascular dementia cases, we found that this observed increase in risk was roughly double that for individuals diagnosed with the less common form of vascular dementia (∼32%) compared to Alzheimer’s disease (∼17%). Together, these findings suggest that shared genetic factors may underlie the development of both coronary artery disease and dementia cases that possess an underlying vascular pathology, perhaps through joint detrimental effects on the vasculature that impact perfusion within both the heart and brain.

Given that an individual’s genotype is fixed at conception, the question of whether this increased genetic risk is deterministic – or is instead mediated through other downstream pathways potentially amenable to modification – is an important consideration for population health and prevention strategies. It is now well-established that much of the genetic risk for CAD can be offset through the adoption of healthy lifestyle behaviours (36–38), and recent years have seen an increased focus on whether the same may be true for dementia (7,8,34). This interest has been spurred on in large part by the 2020 Lancet Commission Report on Dementia, in which it has been estimated that up to 40% of all dementias may be preventable by addressing twelve potentially modifiable risk factors with well-established links to heart disease (39). In support of these claims, we found that risk for incident all-cause dementia over 14 years of follow-up was lower in those with better cardiovascular health scores at their baseline assessment, regardless of underlying genetic risk. Furthermore, this reduction appeared to be driven almost exclusively through a reduced incidence of vascular dementia diagnoses, as well as being broadly mirrored by the magnitude of white matter hyperintensity burden measured in a subset of participants undergoing neuroimaging assessments. Together, these findings have several potentially important implications. First, with the heritability of Alzheimer’s disease predicted to be in the range of 70-80% (40), opportunities to modify AD-related pathologies such as amyloid and tau accumulation may be challenging except through pharmacological means. Our findings instead support a role for improved vascular health as one of the key targets for future dementia prevention strategies. Indeed, the improved management of cardiovascular disease in recent decades is hypothesized to provide the most likely explanation for the unexpected reductions in age-standardized dementia rates that have recently been observed in the developed world (41). It should be noted that these improvements are commonly believed to occur not only through a reduction in overt vascular dementia cases, but by also increasing the resilience of the brain to the negative effects of AD-specific pathologies encountered in Alzheimer’s disease (42). While we saw evidence here of the former, we did not observe any evidence for a protective association between cardiovascular health and incidence of Alzheimer’s disease – findings which contrast with other studies from this cohort (7,34). Reasons for these discrepancies are unclear but may involve the choice of the LE8 cardiovascular health score as our exposure, a composite score which contains a range of both behavioral and biological risk factors which may not all be specific or relevant to Alzheimer’s disease. Second, when assessing underlying neuroimaging phenotypes in a subset of the cohort recalled for MRI brain imaging, we observed significantly higher volumes of white matter hyperintensities (a well-established marker of vascular damage) in individuals at increased genetic and lifestyle risk for CAD. These associations appeared to be additive, such that those at high risk for CAD through both genes and lifestyle were found to have WMH volumes ∼30% higher than those at low risk. In contrast, cortical grey matter volumes and hippocampal volumes – two imaging metrics with well-established links to Alzheimer’s disease – showed no association with our CAD PRS and demonstrated only negligible associations with our lifestyle risk score. These findings again suggest that the biological pathways underlying the observed associations in this study are likely to at least in part differ from those commonly studied in relation to Alzheimer’s disease, and instead may act through pathways predominantly compromising the cerebrovasculature.

Future work is required to investigate these biological pathways and to determine how they link genetic risk for CAD to late-life dementia incidence. Recent work using Mendelian Randomization techniques to investigate a potential causal connection between these diseases has found little evidence connecting manifest CAD to brain health (43), suggesting that overt ischemic heart disease *per se* is unlikely to be the direct causal mechanism influencing dementia risk. The use of a genome-wide PRS in the current study instead suggests that both CAD and vascular dementia may instead possess a shared underlying genetic architecture that simultaneously increases risk of both diseases. This risk could manifest through the downstream mediation of established biological risk factors lying on the causal pathway between genes and both diseases (i.e. vertical pleiotropy), as has been suggested in one recent study for blood pressure (44). Alternatively, associations could represent evidence of gene-environment correlations (where genes influence an individual’s exposure to adverse social environments that themselves influence risk), or horizontal pleiotropy (where CAD genes influence dementia risk through other traits independent of CAD *per se*). Regardless of the underlying mechanisms, however, our findings demonstrate that genetic risk for CAD also increases risk of vascular dementia, particularly in those who do not adhere to well-established cardiovascular health guidelines earlier in their lifecourse.

This study has a number of limitations. First, participants reporting non-White ancestry were excluded to minimise residual confounding due to population stratification. Future studies addressing this research question in diverse ancestries are therefore warranted. Second, information about some lifestyle factors were obtained by self-report and biases may inevitably occur, as opposed to obtaining objective data where possible. Third, healthy lifestyle factors were not randomly assigned, unlike genetic makeup. Thus, despite efforts to adjust for a number of potentially confounding factors, it is possible that genetics or other external factors were responsible for the relationship between lifestyle risk and dementia. Finally, dementia diagnoses were obtained from electronic health records, which in validation studies have been shown to have a lower positive predictive value for vascular dementia compared to all-cause dementia and Alzheimer’s disease (21).

In conclusion, in this large prospective UK population-based cohort, we provide evidence for an association between an elevated genetic risk for coronary artery disease and an increased risk of dementia. Importantly, this risk was reduced in individuals maintaining optimal cardiovascular health in the years preceding diagnosis, predominantly through reductions in incident vascular dementia. Overall, our findings indicate that, regardless of underlying genetic risk for CAD, lifestyle modifications targeting cardiovascular risk may also lower risk of dementia in older age, especially in diagnoses with an underlying vascular component.

## Data Availability

All data used in this publication are open access and available to bona fide researchers through well-documented processes detailed at https://www.ukbiobank.ac.uk/. The statistical code for all analyses in this paper can be found in an open-access GitHub repository located at https://github.com/scottchiesa/UKB_PRS_LRS_Dementia.

https://www.ukbiobank.ac.uk/

## ACKNOWLEDGEMENTS

This research has been conducted using the UK Biobank Resource under Application Number 71702 and includes data provided by patients and collected by the NHS as part of their care and support.

## ABBREVIATIONS

BMI: Body mass index
BP: Blood pressure
CAD: Coronary artery disease
CI: Confidence interval
CVD: Cardiovascular disease
HbA1c: Haemoglobin A1c
HR: Hazard ratio
ICD: International Classification of Diseases
LDL-C: Low-density lipoprotein cholesterol
LE8: Life’s Essential 8
LRS: Lifestyle risk score
PRS: Polygenic risk score
SD: Standard deviation

